# National, state, and county estimates of adult overweight and obesity from electronic health records and kiosks in retail locations, 2024-2025

**DOI:** 10.1101/2025.08.09.25333364

**Authors:** Jithin Sam Varghese

## Abstract

**Importance:** High body mass index (BMI) is a leading cause of morbidity and mortality from non-communicable diseases and increases susceptibility to infectious diseases. Inadequate subnational geographical resolution, low response rates (23-45%), and lag of 2-3 years requires the evaluation of alternative data sources for monitoring the burden of high BMI.

**Objective:** To describe the prevalence of overweight and obesity among individuals aged 18 years and older using electronic health records and kiosks in retail locations.

**Design:** Cross-sectional analysis, January 2024-July 2025

**Setting:** 3,144 counties from 50 states and District of Columbia

**Participants:** 85 million patients from Epic Cosmos and 1.2 million users of Pursuant Health kiosks

**Exposures:** National, state, and county of residence; age, gender, race and ethnicity, urban vs rural residence

**Main Outcomes and Measures:** Overweight (BMI: 25-29.9 kg/m^2^) or obesity (BMI ≥30 kg/m^2^) were defined using height (Epic Cosmos: measured, kiosks: self-reported) and measured weight based on World Health Organization cutoffs. Prevalence was estimated directly for Epic Cosmos and using multilevel regression & post-stratification based on socio-demographic variables for kiosks. We compared these estimates with those from the National Health and Nutrition Examination Survey (NHANES) and Behavioral Risk Factor Surveillance System (BRFSS). Modeled county estimates were compared to PLACES 2024 small area estimates, based on BRFSS 2022, using Local Moran’s I and Spearman rank correlations.

**Results:** The analytic sample was 32.3 % aged 45 to 64 years, 30.9% aged 65 years and older, 55.8% women and 2.6% rural for Epic Cosmos. The analytic samples were 35.8% aged 45 to 64 years, 25.2% aged 65 years and older, 43.7% women and 18.2% rural for kiosks. The prevalence of overweight was 39.1% and 33.6%, and obesity was 43.7% and 43.3% in Epic Cosmos and kiosks respectively, and similar to NHANES and BRFSS. Higher prevalence was observed among NH Black adults, those aged 45 to 64 years, and rural residents for both Epic Cosmos and kiosks. County hotspots were observed across Southern and Midwestern states, and were correlated with PLACES (Cosmos = 0.64, kiosks: 0.34).

**Conclusions and Relevance:** In this administrative EHR and convenience kiosk samples of US adults, prevalence of overweight and obesity were high. National surveys mask substantial subnational variations in prevalence of high BMI.

**KEY POINTS:** *Question:* What is the prevalence of overweight and obesity in states and counties from samples of US adults based on electronic health records and users of health kiosks in retail stores?

*Findings:* Of 89 million patients in Epic Cosmos and 1.2 million users of Pursuant Health kiosks between 2024-2025, nearly three out of four had overweight or obesity. Prevalence was higher among rural and racial and ethnic minorities, as well as in counties in the South and Midwest.

*Meaning:* The increasing availability of data in administrative and convenience samples enable spatially granular and timely population health monitoring of the prevalence of overweight and obesity. These samples cover demographic groups traditionally underrepresented in national surveys.

## Introduction

High body mass index (BMI) is a major risk factor for morbidity and mortality from non-communicable diseases like hypertension, diabetes, atherosclerotic cardiovascular disease, heart failure, and some cancers, and increases susceptibility to infectious diseases. Nearly half of US adults have obesity, defined as BMI of 30 kg/m^2^ or greater, with all states having a prevalence of 20% of more.^1,2^ Accurate population health monitoring of overweight and obesity can inform federal and state public health officials on prioritizing geographic areas and population groups for lifestyle and pharmacological interventions to reduce the disease burden. Although monitoring of overweight and obesity have traditionally relied on sample surveys like the National Health and Nutrition Examination Survey (NHANES) and Behavioral Risk Factor Surveillance System (BRFSS) conducted by the Centers for Disease Control and Prevention, these surveys are limited in their spatial resolution, have declining response rates, and are often lagged by two to three years. Therefore, a critical need exists to evaluate alternative data sources for population health monitoring of overweight and obesity.

The wide adoption of electronic health record systems across national health systems, and technological advancements related to remote monitoring have led to a rapid increase in availability and consideration of novel data sources for monitoring the prevalence of overweight and obesity in the United States.^3,4^ Administrative data, such as electronic health records (EHR) or claims, are collected for purposes other than population health monitoring, but most in-person visits to a hospital involve the measurement of height and weight. However, inclusion in these samples are contingent on healthcare access, which may still exclude socio-economically vulnerable populations. Health kiosks located in convenient retail locations are an example of convenience samples. Kiosks may be able to overcome limitations related to insurance or cost and enable monitoring among these underserved populations. Nevertheless, both administrative and convenience samples are susceptible to several biases. For instance, both convenience samples and administrative data are not based on a probability model for participant selection, with the latter relying heavily on ease of participation. However, appropriate specifications of statistical models may be able to mitigate the selection biases associated with using these data sources for public health.

Our objective is to estimate the prevalence of overweight and obesity in states and counties from an administrative (Epic Cosmos EHRs) and convenience sample (Pursuant Health kiosks) and compare them with national surveys. In an era of declining survey response, findings from this study can inform the value in adoption of alternative data sources for health monitoring, planning or evaluating interventions and policymaking.^3,4^

## Methods

### Study Population

We used cross-sectional data of non-pregnant adults (≥18 years) living in the United States from two probability samples and two non-probability samples for this study. National Health and Nutrition Examination Survey (NHANES) is a nationally representative multistage sample survey of non-institutionalized, community dwelling adults conducted by the National Center for Health Statistics (NCHS). NHANES comprises interviews, physical and laboratory examinations. The 2021-2023 cycle of NHANES had a response rate of 34.5% for the interview and 25.6% for the examination.^5^ The analytic sample for NHANES consisted of 8860 adults.

The Behavioral Risk Factor Surveillance System is a state-based representative survey of over 400,000 adults that is conducted each year in all 50 states, District of Columbia, Guam, Puerto Rico, and the Virgin Islands. We excluded participants from the territories for this analysis. BRFSS is a telephone-based (both cellular and landline) survey, and hence all outcomes are based on self-report. The 2022 cycle of BRFSS had a response rate of 45.0% (range for states: 22.8-66.8%).^6^ The analytic sample for BRFSS consisted of 442,821 adults.

Epic Cosmos Research Platform integrates HIPAA-complaint, deidentified data from over 300 million patients seen at healthcare organizations that use the Epic EHR.^7^ This study used EHRs of patients who used an Epic Cosmos participating institution and lived in one of the 50 states or Washington DC between 1 January 2024 and 10 July 2024. Among 125,646,543 non-pregnant adult patients meeting the inclusion criteria in Epic Cosmos, only 89,081,708 (70.6%) had a valid BMI measurement and were included in the study.

Pursuant Health provides free screenings of blood pressure and weight in Walmart and CVS through over 4,600 voluntary self-service kiosks.^8^ The kiosks are classified by the Food and Drug Administration (FDA) as Class II Medical Devices. Session data was provided as a Limited Data Set as described under the HIPAA Privacy Rule after a data use agreement (DUA) approved by authors (LF and CG) who were not involved the data analysis and interpretation. The sample for Pursuant Health (“kiosks”) consisted of 442,821 non-pregnant adults whose first session where BMI was measured in the calendar period was selected for this analysis. A flowchart for selection of the analytic sample for different data sources is provided in **Supplementary Figure 1**.

### Data collection and variable specification

#### Overweight and Obesity

The method of measurement and calculation of height and weight varied by data source. Weight and height were assessed using standardized protocols for NHANES, self-reported for BRFSS, and extracted from EHRs for Cosmos. Height was self-reported and weight was measured at the kiosk based on standardized instructions. Overweight and obesity were defined as BMI between 25 to 29.9 kg/m^2^ and 30 kg/m^2^ or greater respectively.

#### Socio-demographic characteristics

Demographic characteristics of respondents (for NHANES and BRFSS), patients (for Cosmos), and users (for kiosks) were extracted from their respective data sources: age (18-19, 20-44, 45-64, 65 and over), non-distinguished sex and gender (male, female), race & ethnicity (Hispanic, Non-Hispanic White, NH Black, NH Asian, NH Other). Residence was classified into urban or rural based on Rural-Urban Commuting Area 2010 codes (urban: 1-9, rural: 10).^9^ We additionally extracted covariates from the 5-year American Community Survey (ACS) 2018-2022 based on a previous study of county-level risk factors associated with cardiovascular disease, namely median household income (in current United States dollars), and percentages of residents unemployed, without a high school education, and without health insurance.^10^

### Statistical Analysis

#### Probability Samples

We used the mobile examination survey weights as provided by NCHS to calculate descriptive characteristics of the analytic sample at the national level for NHANES, and the proportion with overweight only, and with obesity. We used the survey weights provided by BRFSS to calculate the national and state estimates. We additionally extracted two sets of estimates derived using BRFSS. First, we extracted projected estimates of overweight and obesity for states for 2024 from the Global Burden of Disease (GBD) Study.^11^ Second, we extracted small area estimates of obesity for counties from PLACES 2024: Local Data for Better Health based on BRFSS 2022, which are released by the CDC.^12^

#### Non-Probability Samples

We calculated direct estimates for states and counties based on counts of individuals with overweight and obesity relative to the analytic sample for Epic Cosmos and kiosks. We also used multilevel regression and post-stratification (MRP) to derive modeled estimates for states and counties. MRP is a two-stage approach of fitting a logistic generalized linear mixed model, followed by using this model to generate estimates and 95% confidence intervals using post-stratification weights based on the ACS.^13^ The logistic mixed model was fit using fixed effects for socio-demographic characteristics (age, sex, race & ethnicity, residence), county characteristics from ACS, and random intercepts for states and counties. A detailed discussion of the statistical methods is reported elsewhere.

#### Comparison of Estimates

We compared the weighted estimates from BRFSS 2022, and modeled estimates from GBD 2024 and PLACES 2024 with the estimates from Epic Cosmos and kiosks using rank correlations. We calculated Local Moran’s I to measure spatial similarity for county estimates from PLACES, Epic Cosmos, and kiosks. All analysis were carried out using Python 3.11 and R 4.3.3.

## Results

The analytic sample of 85 million adults in Epic Cosmos and 1.2 million adults in Epic Cosmos covered 50 states and Washington DC. The sample was 32.3 % aged 45 to 64 years, 30.9% aged 65 years and older, 55.8% women and 2.6% rural for Epic Cosmos. The sample was 35.8% aged 45 to 64 years, 25.2% aged 65 years and older, 43.7% women and 18.2% rural for kiosk users.

The distribution of age and sex as well as mean height, weight, and BMI were similar to the weighted estimates from NHANES and BRFSS (**Table 1**). The racial and ethnic distribution was similar to NHANES, BRFSS for Epic Cosmos but was disproportionately Hispanic adults among kiosk users. The unweighted prevalence of obesity was higher in Epic Cosmos (43.7%) and Pursuant (42.5%), relative to estimates from NHANES (39.5%) and BRFSS (34.1%)

**Table 1.** Comparison of demographic characteristics of kiosk users with national samples.

|  | **NHANES 2021-2023<sup>a</sup>** | **BRFSS 2022<sup>a</sup>** | **Epic Cosmos 2024-2025<sup>b</sup>** | **Pursuant 2024-2025<sup>b</sup>** |
| --- | --- | --- | --- | --- |
| N (Unweighted) | 6194 | 387,549 | 85,186,060 | 1,180,273 |
| Age category (%) |  |  |  |  |
| 18 to 19 | 3.4 (2.7, 4.4) | 3.5 (3.3, 3.6) | 2.8 | 1.4 |
| 20 to 44 | 42.7 (40.0, 45.4) | 41.4 (41.1, 41.7) | 34 | 37.7 |
| 45 to 64 | 32.2 (29.8, 34.6) | 31.5 (31.2, 31.8) | 32.3 | 35.8 |
| 65 plus | 21.7 (19.4, 24.2) | 23.6 (23.4, 23.9) | 30.9 | 25.2 |
| Men (%) | 49.1 (47.7, 50.5) | 49.3 (48.9, 49.6) | 44.2 | 56.3 |
| Women (%) | 50.9 (49.5, 52.3) | 50.7 (50.4, 51.1) | 55.8 | 43.7 |
| Race and Ethnicity (%) |  |  |  |  |
| Hispanic | 16.8 (11.9, 23.2) | 16.2 (15.9, 16.5) | 12.5 | 22.3 |
| Non-Hispanic White | 59.9 (55.8, 63.9) | 59.1 (58.7, 59.4) | 60.5 | 48.2 |
| Non-Hispanic Black | 10.7 (8.26, 13.8) | 11.5 (11.3, 11.7) | 13.2 | 13.4 |
| Non-Hispanic Other |  | 7.4 (7.2, 7.6) | 9.9 | 10.3 |
| Non-Hispanic Asian | 12.5 (10.4, 15.0) | 5.9 (5.7, 6.1) | 3.9 | 5.7 |
| Rural residence (%) | - | 6.4 (6.3, 6.5) | 2.6 | 18.2 |
| Height (cm) | 168.0 (167.6, 168.5) | 170.4 (170.3, 170.5) | 170.2 | 170.0 |
| Weight (kg) | 83.6 (82.1, 85.2) | 82.9 (82.7, 83.0) | 85.3 | 86.4 |
| Body Mass Index (kg/m<sup>2</sup>) | 29.6 (29.0, 30.1) | 28.5 (28.4, 28.5) | 30.5 | 29.7 |
| <18.5 kg/m<sup>2</sup> | 1.8 (1.5, 2.3) | 2.0 (1.9, 2.1) | 2.7 | 2.5 |
| 18.5-24.9 kg/m<sup>2</sup> | 26.9 (23.9, 30.1) | 30.5 (30.2, 30.9) | 14.5 | 23.0 |
| 25-29.9 kg/m<sup>2</sup> | 31.7 (30.1, 33.5) | 33.3 (33.0, 33.6) | 39.1 | 32.0 |
| ≥30 kg/m<sup>2</sup> | 39.5 (35.7, 43.5) | 34.1 (33.8, 34.4) | 43.7 | 42.5 |

a Weighted estimates as percentages with 95% CI or mean (95% CI for survey weighted analyses)
b Direct estimates as crude percentages (%) or mean.
Pursuant 2024-2025: 1,180,273 (19,802 excluded for missing race-ethnicity before modeling, 1 excluded for missing gender)

Prevalence of overweight and obesity varied between socio-demographic groups with highest prevalence of obesity among women, those aged 45-64 years, Non-Hispanic Blacks, and rural residents in both Epic Cosmos and kiosk samples (**Table 2**). Modeled estimates of overweight and obesity were higher than unweighted estimates. Consistent with national estimates, prevalence of obesity was higher for Epic Cosmos and kiosk users, relative to NHANES and BRFSS for most population groups (**Supplementary Table 1**). For example, among women, prevalence of obesity was 44.5% in Epic Cosmos and 46.1% (95%CI: 44.9-47.3) among kiosk users but 40.9% (95%CI: 36.4-45.4) in NHANES and 34.1% (95%CI: 33.6-34.5) in BRFSS.

**Table 2.** Prevalence of overweight and obesity.

|  | **Epic Cosmos 2024-2025** |  |  | **Pursuant 2024-2025 (Direct)** |  |  | **Pursuant 2024-2025 (Modeled)** |  |
| --- | --- | --- | --- | --- | --- | --- | --- | --- |
|  | **N** | **Overweight** | **Obesity** | **N** | **Overweight** | **Obesity** | **Overweight** | **Obesity** |
| Total | 85186060 | 39.1 | 43.7 | 1,180,273 | 32.0 | 42.5 | 33.6 (32.8, 34.4) | 43.3 (42.3, 44.4) |
| Gender |  |  |  |  |  |  |  |  |
| Women | 47495167 | 35.6 | 44.5 | 515,546 | 26.8 | 47.8 | 30.0 (29.1, 31.0) | 46.1 (44.9, 47.3) |
| Men | 37663989 | 43.6 | 42.8 | 664,727 | 36.0 | 38.4 | 37.2 (36.2, 38.3) | 40.5 (39.3, 41.7) |
| Other/Unknown | 26904 | 33.8 | 37.1 |  |  |  |  |  |
| Age category |  |  |  |  |  |  |  |  |
| 18-19 | 2382118 | 25.7 | 21.7 | 15,936 | 24.6 | 27.4 | 28.2 (27.1, 29.4) | 29.2 (27.9, 30.3) |
| 20-44 | 28953255 | 34.7 | 41.9 | 445,071 | 28.1 | 45.8 | 31 (29.9, 32.2) | 45.7 (44.2, 47.1) |
| 45-64 | 27493317 | 39.5 | 50.9 | 422,025 | 31.9 | 47.3 | 34.3 (33.1, 35.4) | 47.3 (45.9, 48.6) |
| 65 and above | 26357370 | 44.9 | 40.2 | 297,241 | 38.3 | 31.6 | 38.5 (37.2, 39.7) | 34.9 (33.6, 36.3) |
| Race & Ethnicity <sup>a</sup> |  |  |  |  |  |  |  |  |
| Hispanic | 11740480 | 40.5 | 48.4 | 263,610 | 32.0 | 47.8 | 34.5 (32.3, 36.7) | 47.8 (45.1, 50.2) |
| Non-Hispanic White | 56808528 | 39.9 | 42.9 | 568,998 | 32.2 | 41.2 | 33.4 (32.6, 34.2) | 42.8 (41.6, 43.9) |
| Non-Hispanic Black | 12436957 | 34.8 | 52.6 | 158,589 | 28.9 | 49.7 | 31.2 (29.9, 32.5) | 50 (48.5, 51.6) |
| Non-Hispanic Asian | 3667798 | 40.4 | 20.6 | 67,260 | 37.5 | 19.0 | 37.9 (35.4, 40.1) | 23.1 (20.9, 25) |
| Non-Hispanic Other | 9314418 | 39.1 | 41.1 | 121,816 | 31.9 | 41.0 | 32.6 (31.4, 33.9) | 44.0 (42.4, 45.5) |
| **Residence** |  |  |  |  |  |  |  |  |
| Urban | 82499692 | 39.1 | 43.6 | 960,842 | 32.3 | 41.6 | 33.8 (32.9, 34.8) | 42.9 (41.6, 44.1) |
| Rural | 2226613 | 37.9 | 48.7 | 214,240 | 30.4 | 46.5 | 32.2 (31.6, 32.9) | 46.4 (45.4, 47.3) |
| Unknown | 459755 | 43.4 | 39 | - | - | - | - | - |

<sup>a</sup> More than one race-ethnicity counted in all applicable categories due to data structure for Epic Cosmos. Therefore, the sum of all categories exceeds total sample size.
Values are crude counts and percentages for Epic Cosmos and Pursuant 2024-2025 (Direct) estimates, and modeled percentages and 95% confidence intervals.

Among those aged 45 to 64 years, prevalence of obesity was 50.9% and 47.3% (95%CI: 45.9-48.6) for Epic Cosmos and kiosk users, but 46.4% (95%CI: 43.0-49.8) and 38.6% (38.1-39.2) in NHANES and BRFSS respectively. Among rural adults, prevalence of obesity was 48.7% in Epic Cosmos and 46.4% (95%CI: 45.4-47.3) among kiosk users but only 38.3% (95%CI: 37.3-39.3) in BRFSS.

Prevalence of overweight (**Figure 1A-C**)and obesity (**Figure 1D-F**) varied between states for BRFSS, Epic Cosmos, and kiosks. States with high burden of obesity also tend to have slightly lower burdens of overweight, suggesting a high population burden of high BMI. States in the South and Midwest, having higher prevalence than states in West and Northeast for all data sources. Prevalence of obesity was highest for Mississippi and West Virginia in Epic Cosmos. Prevalence of overweight was more homogeneous across states for BRFSS, Epic Cosmos and kiosks, and correlations of overweight prevalence between data sources weaker than those of obesity. Prevalence of overweight or obesity for states was also correlated with projections from the GBD study for 2024 for Epic Cosmos and kiosks (**Supplementary Figure 2**).

**Figure 1.**
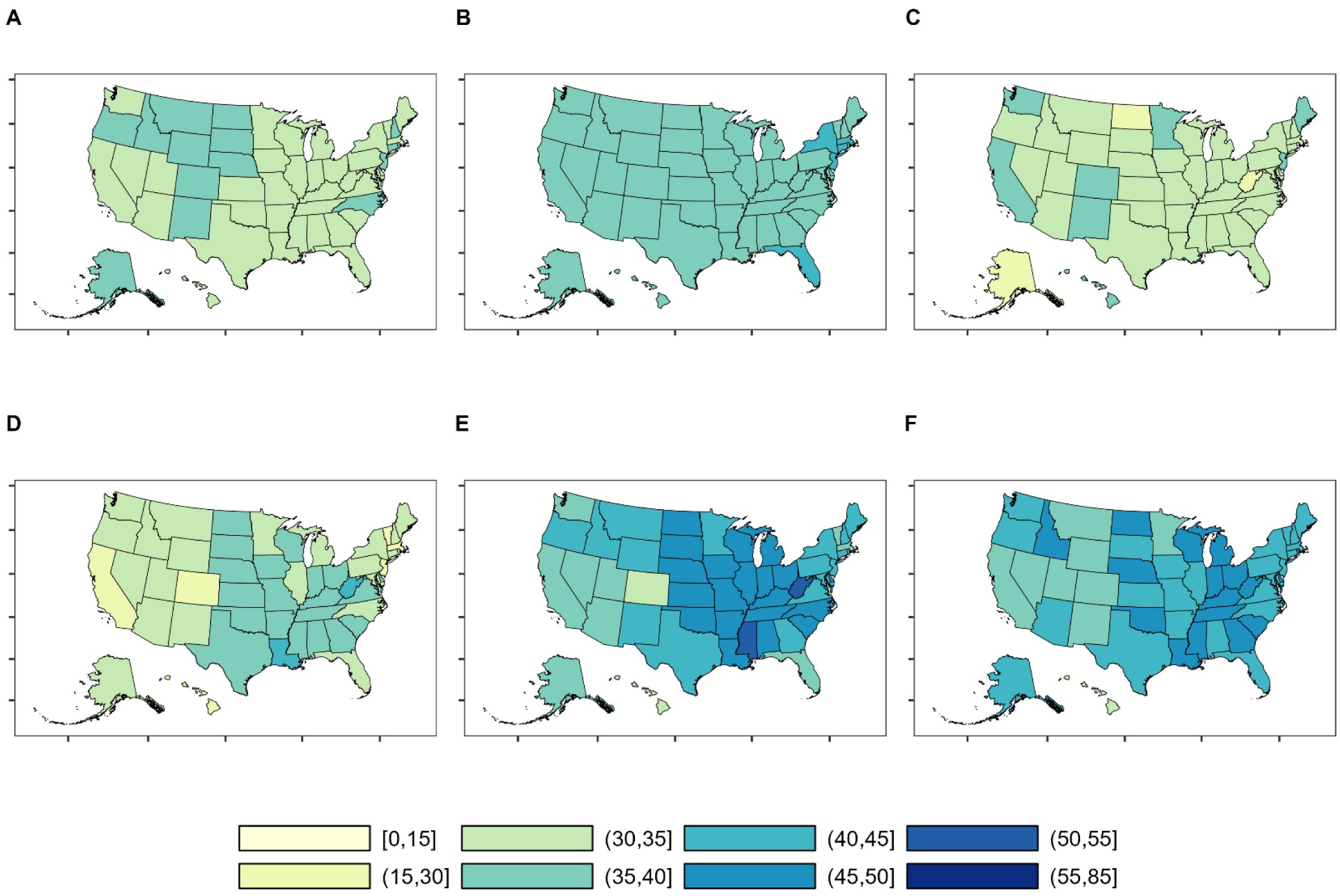
State prevalence of overweight and obesity. All estimates are prevalence based on weighted estimates from BRFSS 2022, unweighted counts in Cosmos and multilevel regression and post-stratification for Pursuant Health kiosks We used World Health Organization cutoffs for overweight (BMI: 25.0-29.9kg/m^2^) and obesity (BMI ≥ 30kg/m^2^). A: BRFSS overweight, B: Cosmos overweight, C: Pursuant overweight, D: BRFSS obesity, E: Cosmos obesity, F: Pursuant obesity

The prevalence of overweight or obesity in counties across the US (**Figure 2**) suggested hotspots for Epic Cosmos (overweight: 0.38, obesity: 0.51), but less for modeled estimates from kiosk users (overweight: 0.05, obesity: 0.14). We observed clustering of high prevalence of overweight for counties in New Mexico and Idaho in both data sources. Nevertheless, estimates were correlated with modeled estimates from PLACES 2024 for both Epic Cosmos (ρ: 0.64 [95%CI: 0.63-0.67]) and kiosks (ρ: 0.34 [95%CI: 0.31-0.37]) (**Figure 3**). Spatial autocorrelation using Local Moran’s I also indicated a similar clustering of low prevalence counties and high prevalence counties based on both Cosmos and kiosks relative to modeled estimates from PLACES 2024 (**Supplementary Table 2**).

**Figure 2.**
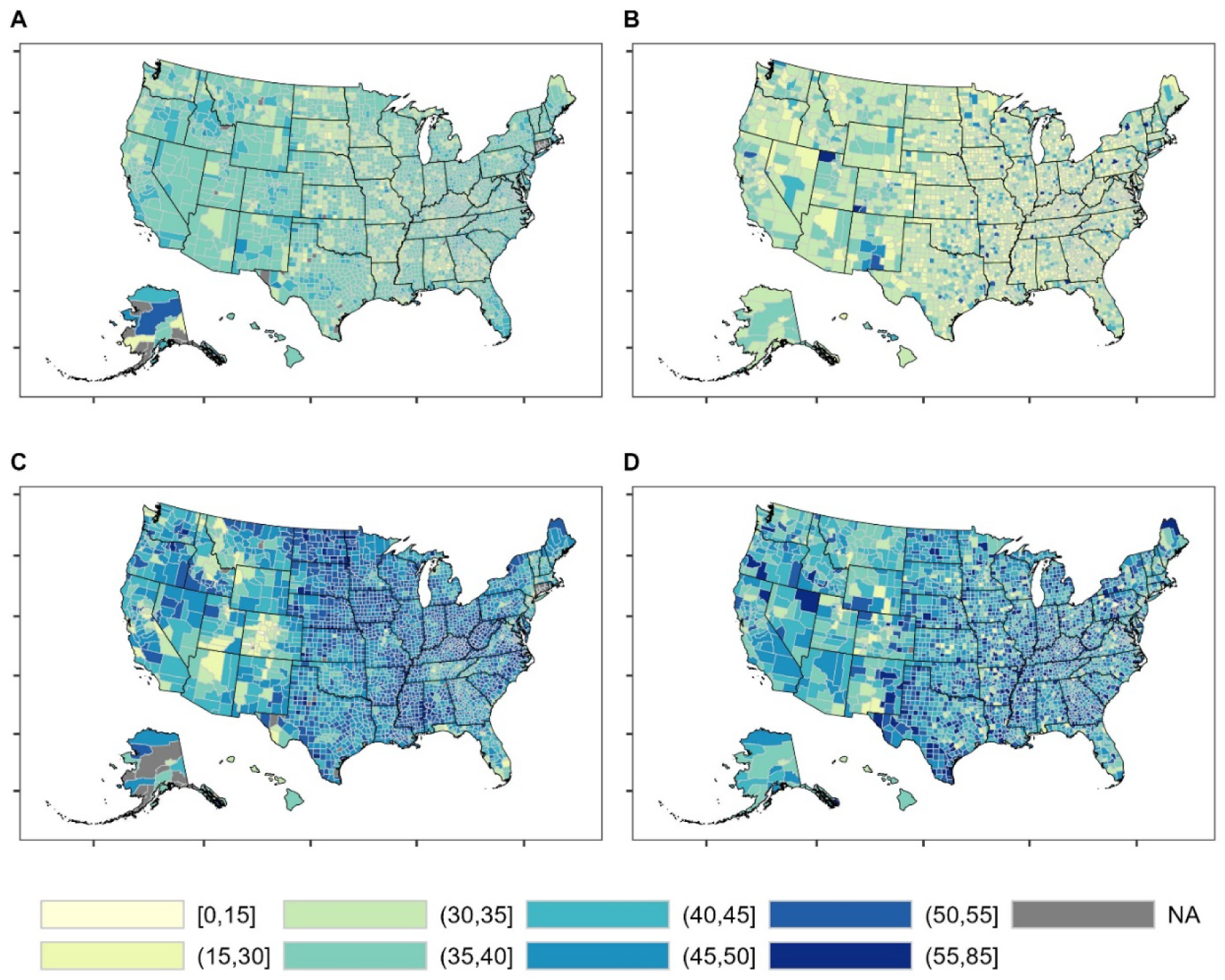
County prevalence of overweight and obesity. All estimates are prevalence based on unweighted counts in Cosmos and multilevel regression and post-stratification for Pursuant Health kiosks We used World Health Organization cutoffs for overweight (BMI: 25.0-29.9kg/m^2^) and obesity (BMI ≥ 30kg/m^2^). A: Cosmos overweight, B: Pursuant overweight, C: Cosmos obesity, D: Pursuant obesity. Standard errors for Pursuant are visualized in **Supplementary Figure 3**. Global Moran’ I for each are 0.38, 0.05, 0.51, and 0.14 respectively.

**Figure 3.**
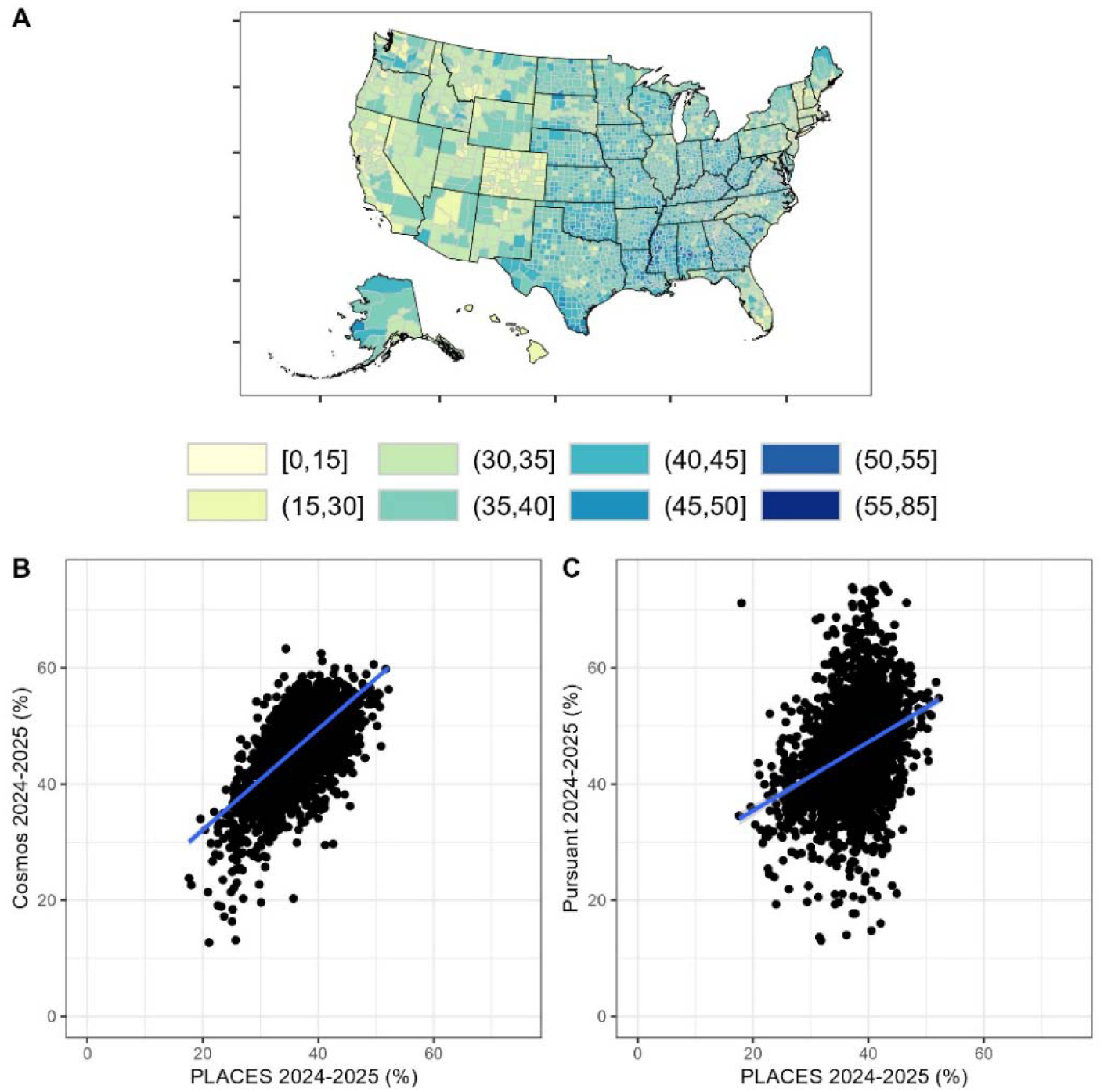
Correlation of obesity between data sources. A: Prevalence of obesity from PLACES 2024 (derived from BRFSS 2022). Panels B and C are scatterplots of PLACES 2024 against Cosmos 2024-2025 (Spearman’s ρ: 0.64 [95%CI: 0.63-0.67]) and Pursuant Health 2024-2025 (Spearman’s ρ: 0.34 [95%CI: 0.31-0.37]). Standard errors for PLACES are visualized in **Supplementary Figure 3**. Global Moran’s I for PLACES 2024 is 0.59.

## Discussion

Overweight or obesity affect over three out of four Americans today, with an additional 42 million adults projected to be added by 2050.^11^ We observed that state and county prevalence of obesity from BRFSS are correlated with estimates from EHRs in Epic Cosmos and users of Pursuant Health kiosks. Furthermore, prevalence of overweight and obesity varied between and within states, suggesting opportunities for targeted weight loss interventions. We also observed a higher prevalence of obesity in the Southern and Midwestern, relative to the Northeastern and Western states, as well as among Hispanic and Non-Hispanic Black populations are consistent with previous studies.^2,14^

Findings from this study suggest that administrative and convenience samples can complement national surveys which are limited in their spatial resolution and have declining response rates. NHANES is nationally representative but response rates for its biennial cycles gradually declined from >75% in 1999-2000 to 25% in 2021-2023 for examinations. Similarly, BRFSS provides precise estimates for states but are based on self-reported data, with similar declining trends in response rates over the last three decades. Although a lower response rate does not indicate bias in estimates,^15^ others have noted response rates for population surveys tend to be lower among low income, rural, and racial & ethnic minority groups.^16,17^ Similar to this study that used MRP to account for convenience sampling, PLACES used MRP to meet the needs of county health officials. Small area estimates are based on modeling assumptions and may differ (correlation: 0.28) from direct survey estimates based on a validation study of BRFSS 2011 using data from Missouri.^18,19^ However, the observed rank and spatial correlation of modeled estimates from PLACES with direct estimates from Cosmos and modeled estimates from kiosks suggest broader secular geographic trends. Nevertheless, the estimates should be validated using other representative data sources such as All Payer Claims Databases.^20^

Based on a serial cross sectional analysis of NHANES, age-adjusted prevalence of obesity did not change between 2013-2014 and 2021-2023.^1^ However, national surveys are lagged by two to three years, limiting the ability of public health officials to track short-term changes in prevalence of overweight and obesity from wide uptake of pharmacological or lifestyle interventions. Administrative and convenience samples are frequently updated, can track individuals longitudinally, and can complement national surveys in monitoring short term trends.

Of note, the increasing rates of prescription of expensive but effective incretin mimetic drugs, such as semaglutide or tirzepatide, may result in faster declines of overweight and obesity in some counties more than others because of access disparities.^21^ These trends are harder to infer from surveys that do not measure all the requisite variables and lack adequate socio-demographic or spatial resolution.

Strengths of this study include the most recent data (up to July 2024) with detailed characterization of probability surveys, and two large non-probability samples for monitoring the prevalence of adult overweight and obesity. However, this study has some limitations. First, our analysis was restricted to body mass index. We did not use ethnicity-specific cutoffs for overweight and obesity, and did not assess body composition to confirm excess adiposity.^22^ We also did not correct for self-report bias since the objective was not to evaluate the prevalence of severe or Class III obesity (BMI ≥ 40 kg/m^2^). An analysis of BRFSS suggests that 8.8% of adults had severe obesity based on the bias corrected BMI, as opposed to 5.3% based on self-reported BMI.^23^ Second, we did not correct for systematic selection of patients into Epic Cosmos participating institutions. Only half of healthcare systems in the United States use Epic software, and these often tend to be well-resourced academic institutions.^24^

In summary, the high burden and spatial variation in overweight and obesity offer opportunities for targeted pharmacological and lifestyle interventions. Although response rates from probability surveys such as NHANES and BRFSS are declining, administrative and convenience samples can complement monitoring efforts. Future studies should explore which geographic areas and socio-demographic populations are best represented by these novel data sources, and whether they, alone or in combination,^25^ can be used to augment national surveys to generate better estimates of population health indicators.

## Ethics approval and consent to participate

All participants gave written informed consent before participation in NHANES and BRFSS. All kiosk users agreed to the Pursuant Health user agreement. All data in Epic Cosmos are HIPAA compliant and expert determined de-identified. We were exempt from ethical approval for the analysis of de-identified secondary datasets by the Institutional Review Board of Emory University.

## Data availability statement

The code for the analysis is available on https://github.com/chroniq-lab/kiosk-user-patterns. Data from BRFSS, NHANES, PLACES are publicly available. Data from GBD are available from the IHME website. Data from Epic Cosmos are available to employees of participating institutions. Data from Pursuant Health are available at the discretion of the company after signing a data use agreement.

## Consent for publication

Not applicable

## Conflicts of interests

Lauren Fede and Cameron Gocke are employees of Pursuant Health. The authors declare no conflicts of interest.

## Funding

None

## Acknowledgements

We thank Mark Hutcheson (Emory Global Diabetes Research Center) and Rajsekhar Guddneppanavar (Emory Office of Technology Transfer) for their administrative support. We thank David Rosenblatt (Pursuant Health) and Leslie Gerdes-Sommers (Pursuant Health) for their inputs on the manuscript. We thank the Epic Cosmos Research team (Danessa Sandmann, Meghan Howat) for their helpful inputs.

## Sources of Support

None

## Abbreviations

ACS: American Community Survey
BMI: Body mass index
BRFSS: Behavioral Risk Factor Surveillance System
CDC: Centers for Disease Control & Prevention
GBD: Global Burden of Disease study
NCHS: National Center for Health Statistics
NHANES: National Health and Nutrition Examination Surveys

## Notes

### Competing Interest Statement

The authors have declared no competing interest.

### Funding Statement

This study did not receive any funding

### Author Declarations

Ethics committee/IRB of Emory University waived ethical approval for this work

