## Supplementary Material for "National, state, and county estimates of adult overweight and obesity from electronic health records and kiosks in retail locations, 2024-2025"

**Supplementary Table 1. Prevalence of overweight and obesity**

|  | **NHANES 2021-2023 Survey Weighted** | | | **BRFSS 2022 Survey Weighted** | | |
| --- | --- | --- | --- | --- | --- | --- |
|  | **Unweighted N** | **Overweight** | **Obesity** | **Unweighted N** | **Overweight** | **Obesity** |
| **Sex** |  |  |  |  |  |  |
| Women | 3377 | 28.59 (26.77, 30.48) | 40.84 (36.39, 45.44) | 197689 | 29.21 (28.77, 29.65) | 34.04 (33.59, 34.5) |
| Men | 2817 | 35.01 (32.17, 37.96) | 38.66 (34.79, 42.67) | 189860 | 38.88 (38.43, 39.33) | 32.61 (32.18, 33.04) |
| **Age category** |  |  |  |  |  |  |
| 18-19 | 265 | 17.73 (12.46, 24.6) | 25.62 (20.54, 31.46) | 5958 | 23.54 (21.67, 25.51) | 15.51 (14.12, 17) |
| 20-44 | 1939 | 31.67 (28.86, 34.61) | 36.65 (31.43, 42.21) | 108922 | 31.92 (31.41, 32.43) | 32.31 (31.79, 32.83) |
| 45-64 | 2021 | 30.69 (28.65, 32.81) | 46.41 (43.01, 49.85) | 128547 | 35.81 (35.26, 36.37) | 38.61 (38.05, 39.17) |
| 65 and above | 1969 | 35.66 (34.11, 37.25) | 38.26 (35.37, 41.24) | 144122 | 37.25 (36.66, 37.85) | 30.64 (30.08, 31.21) |
| **Race & Ethnicity** |  |  |  |  |  |  |
| Hispanic | 1066 | 33.12 (30.05, 36.33) | 43.04 (35.27, 51.18) | 30972 | 35.74 (34.69, 36.8) | 36.72 (35.65, 37.79) |
| Non-Hispanic White | 3620 | 32.16 (30.19, 34.19) | 39.16 (35.59, 42.84) | 289595 | 34.5 (34.17, 34.84) | 32.47 (32.14, 32.8) |
| Non-Hispanic Black | 751 | 27.67 (24.18, 31.46) | 51.11 (46.38, 55.82) | 29914 | 31.56 (30.58, 32.55) | 43.03 (42.01, 44.05) |
| Non-Hispanic Asian | - | - | - | 10839 | 31.97 (30.29, 33.7) | 13.29 (11.96, 14.75) |
| Non-Hispanic Other | 757 | 31.37 (25.43, 37.99) | 28.54 (21.49, 36.81) | 26229 | 33.13 (31.95, 34.34) | 33.46 (32.27, 34.66) |
| **Residence** |  |  |  |  |  |  |
| Rural | - | - | - | 49782 | 33.71 (32.76, 34.68) | 38.31 (37.33, 39.3) |
| Urban | - | - | - | 337672 | 34.14 (33.81, 34.47) | 32.97 (32.65, 33.3) |
| Unknown | - | - | - | 95 | 42.12 (29.01, 56.45) | 29.27 (18.89, 42.38) |

Non-pregnant adults with BMI measured

**Supplementary Table 2. Spatial autocorrelation measured using Local Moran’s I**

| **PLACES 2024 Obesity** | **Cosmos 2024-2025 Obesity** | | | | **Pursuant 2024-2025 Obesity** | | | |
| --- | --- | --- | --- | --- | --- | --- | --- | --- |
|  | **Low-Low** | **High-Low** | **Low-High** | **High-High** | **Low-Low** | **High-Low** | **Low-High** | **High-High** |
| **Low-Low** | 766 | 116 | 101 | 186 | 620 | 244 | 183 | 146 |
| **High-Low** | 104 | 97 | 37 | 138 | 110 | 94 | 65 | 110 |
| **Low-High** | 108 | 26 | 103 | 157 | 87 | 86 | 123 | 105 |
| **High-High** | 229 | 106 | 114 | 716 | 133 | 198 | 251 | 589 |

**Supplementary Figure 1. Flowchart of participants**

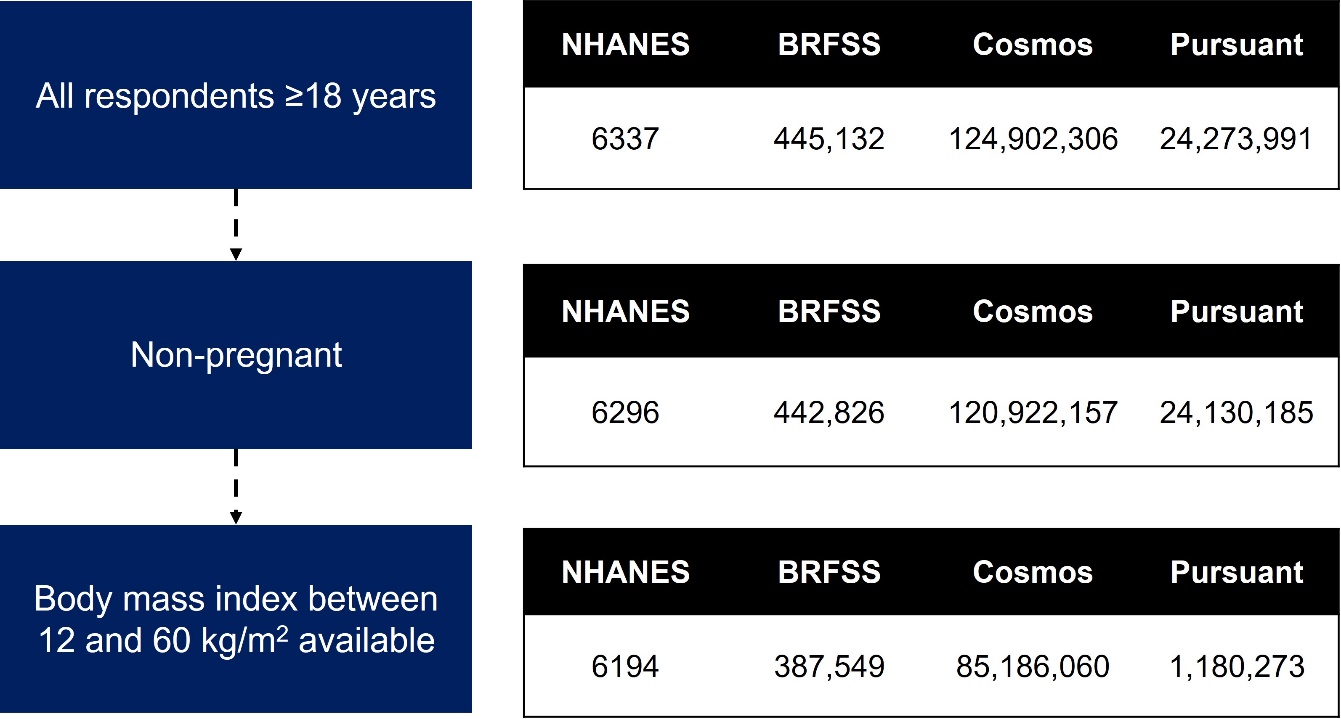

Note: For BRFSS, we additionally excluded participants from Puerto Rico, Virgin Islands, and Guam Removed duplicate sessions at the last step. For Cosmos, patients needed to be from the 50 states or DC in the first step. Pregnancy was excluded based on active sproblem diagnoses which might have low sensitivity. For Pursuant we removed duplicate sessions at the last step.

**Supplementary Figure 2. Correlation of state overweight and obesity prevalence with weighted estimates from BRFSS 2022 and Global Burden of Disease Study 2024 projections**

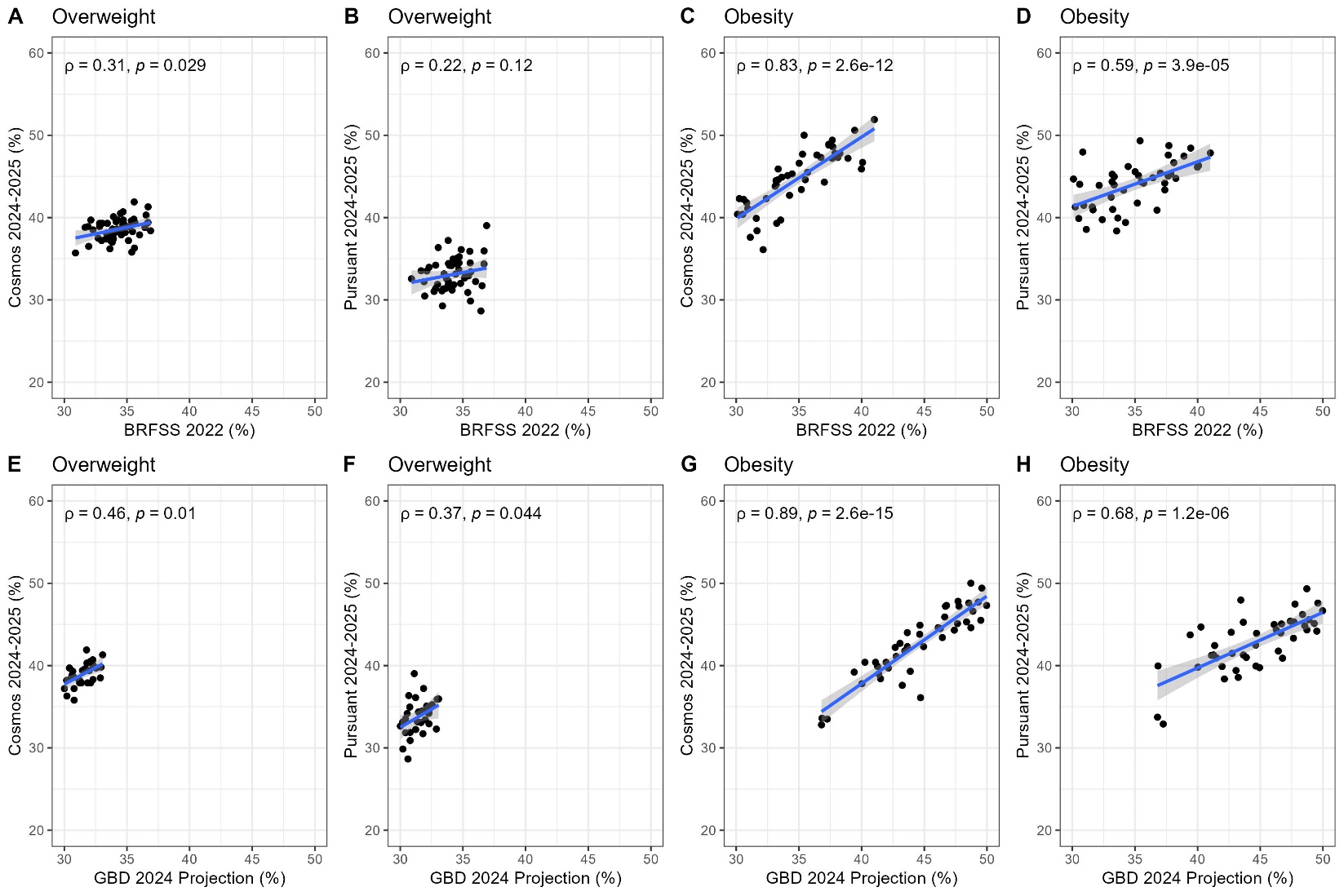

Overweight only: 25-29.9 kg/m^2^; Obesity: ≥30 kg/m^2^

Spearman correlations and two-sided hypothesis test

**Supplementary Figure 3. Precision of modeled estimates from PLACES 2024 and Pursuant Health 2024-2025**

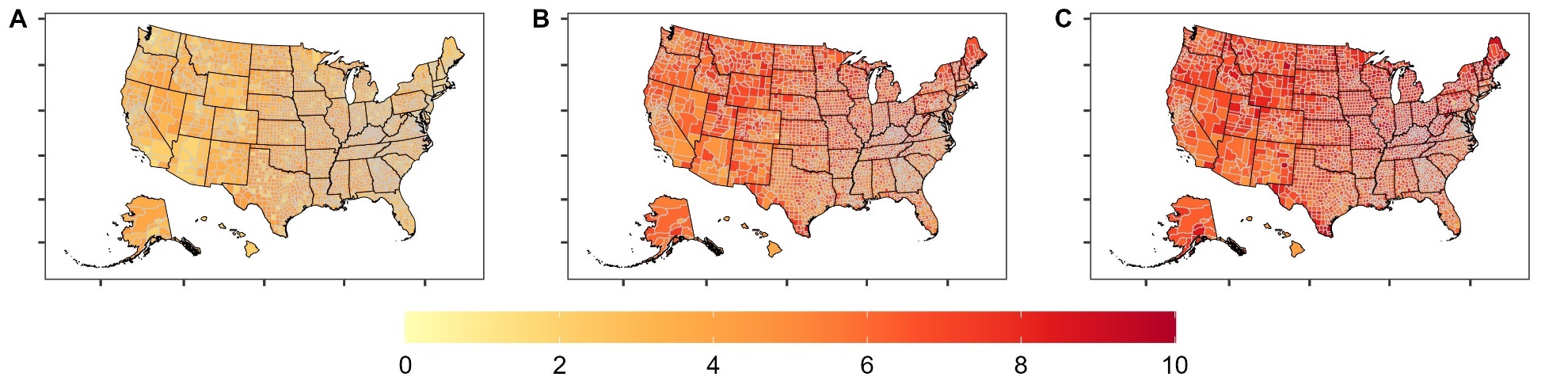

A: PLACES 2024

B: Pursuant Health 2024-2025 Overweight

C: Pursuant Health 2024-2025 Obesity
